# Effectiveness of ChAdOx1-S COVID-19 Booster Vaccination against the Omicron and Delta variants in England

**DOI:** 10.1101/2022.04.29.22274483

**Authors:** Freja Kirsebom, Nick Andrews, Ruchira Sachdeva, Julia Stowe, Mary Ramsay, Jamie Lopez Bernal

## Abstract

**Background:** Despite the potential widespread global use of the ChAdOx1-S booster, to date there are no published data on the real-world effectiveness. VE studies have found one and two doses of the ChAdOx1-S vaccine to be highly effective, and clinical trial data have demonstrated enhanced immunity following a ChAdOx1-S booster. In England, some individuals received a ChAdOx1-S booster where vaccination with mRNA vaccines was clinically contraindicated.

**Methods:** The demographic characteristics of those who received a ChAdOx1-S booster were compared to those who received a BNT162b2 booster. A test-negative case control design was used to estimate vaccine effectiveness of the ChAdOx1-S booster against symptomatic disease and hospitalisation in England.

**Findings:** Those who received a ChAdOx1-S booster were more likely to be female (adjusted odds ratio (OR) 1.67 (1.64-1.71)), in a clinical risk group (adjusted OR 1.58 (1.54-1.63)), in the CEV group (adjusted OR 1.84 (1.79-1.89)) or severely immunosuppressed (adjusted OR 2.05 (1.96-2.13)).

Protection against symptomatic disease in those aged 65 years and older peaked at 66.1% (16.6 to 86.3%) and 68.5% (65.7 to 71.2%) amongst those who received the ChAdOx1-S and BNT162b2 booster vaccines, respectively. Protection waned to 44.5% (22.4 to 60.2%) and 54.1% (50.5 to 57.5%) after 5-9 weeks. Protection against hospitalisation following Omicron infection peaked at 82.3% (64.2 to 91.3%) after receiving a ChAdOx1-S booster, as compared to 90.9% (88.7 to 92.7%) for those who received a BNT162b2 booster.

**Interpretation:** Differences in the population boosted with ChAdOx1-S in England renders direct comparison of vaccine effectiveness by manufacturer challenging. Nonetheless, this study supports the use of the ChAdOx1-S booster for protection against severe disease with COVID-19 in settings that have not yet offered booster doses and suggests that those who received ChAdOx1-S as a booster in England do not require re-vaccination ahead of others.

**Funding:** UKHSA

## Background

Real-world studies on the vaccine effectiveness (VE) and duration of protection conferred by booster vaccines against mild and severe outcomes of COVID-19 infection have primarily focussed on mRNA vaccine boosters (1-4). Over 1.6 billion booster doses have been administered globally, including with the adenoviral vector based ChAdOx1-S (Vaxzevria/Covishield, AstraZeneca) vaccine (5). The ChAdOx1-S booster is in use as part of both homologous and heterologous vaccine programmes in South America, Africa and Asia (6-8), and has been relied upon particularly in low- and middle-income countries due to the lower cost and the relative logistical ease of use as the cold-chain requirements are less extensive than those of the mRNA vaccines.

Real-world VE studies have found one and two doses of the ChAdOx1-S vaccine to be moderately effective against mild disease and highly effective against severe COVID-19, with VE against symptomatic disease peaking around 50% whilst VE against hospitalisation peaks at 85-90% (9-13). Nonetheless, waning over time occurs after a primary course (two doses) (14). Waning against symptomatic disease caused by the Omicron variant occurs from a month after the second dose (9). Protection against more severe outcomes (hospitalisation and death) is maintained for longer, but by 25 or more weeks after a second dose protection against hospitalisation with Omicron is estimated at around 30% (15). Clinical trial data have indicated that ChAdOx1-S is highly immunogenic when given as a booster following primary vaccination (16-18), although to date there are limited data on the real-world effectiveness. Given the potential for widespread use of ChAdOx1-S vaccine as a booster, there is an urgent need to understand the level and duration of protection conferred in real-world settings.

The UK COVID-19 vaccination program has been in place since December 2020 with primary courses of two doses of either BNT162b2 (Pfizer-BioNTech, Comirnaty®), ChAdOx1-S or mRNA-1273 (Spikevax, Moderna). From May 2021, ChAdOx1-S primary immunisation was not recommended as first line vaccination for individuals not in a clinical risk group aged under 40 years following reports of rare cases of concurrent thrombosis and thrombocytopenia after the first ChAdOx1-S dose (19, 20). Booster vaccination with either BNT162b2 or a half dose (50µg) of mRNA-1273 was introduced in September 2021 to adults over 50 years old and those in risk groups, and in November 2021 expanded to all adults. A small number of individuals who had received at least one dose of ChAdOx1-S previously and for whom vaccination with both BNT162b2 and mRNA-1273 were clinically contraindicated, were recommended booster vaccination with the ChAdOx1-S vaccine (21). Clinical contraindications included individuals with a prior allergic reaction to any component of the vaccine e.g. polyethylene glycol, or individuals who had a previous systemic anaphylaxis reaction to a COVID-19 vaccine (21). ChAdOx1-S was in some cases also offered for logistical reasons, for example, to housebound individuals.

In this study we estimate VE against symptomatic disease and hospitalisation following Delta or Omicron infection after booster vaccination in adults in England who received a ChAdOx1-S booster vaccine as compared to those who received a BNT162b2 booster following a ChAdOx1-S primary course.

## Methods

### Study Design

To estimate vaccine effectiveness against symptomatic disease and hospitalisation, a test negative case control design was used. The odds of vaccination in symptomatic PCR positive cases was compared to the odds of vaccination in symptomatic individuals aged 18 years and older who tested negative for SARS-CoV-2 in England.

### Data Sources

#### COVID-19 Testing Data

Data on all positive PCR and LFTs, and on negative Pillar 2 PCR tests from symptomatic individuals with a test date after 25 November 2020 were extracted up to 17 February 2022. Any negative tests taken within 7 days of a previous negative test, and any negative tests where symptom onset date was within the 10 days or a previous symptoms onset date for a negative test were dropped as these likely represent the same episode. Negative tests taken within 21 days of a subsequent positive test were also excluded as chances are high that these are false negatives. Positive and negative tests within 90 days of a previous positive test were also excluded; however, where participants had later positive tests within 14 days of a positive then preference was given to PCR tests and symptomatic tests. For individuals who had more than one negative test, one was selected at random in the study period. Data were restricted to persons who had reported symptoms and gave a symptom onset date within the 10 days before testing to account for reduced PCR sensitivity beyond this period in an infection event.

#### Vaccination Data

To estimate the odds of an individual receiving a ChAdOx1-S booster following a ChAdOx1-S primary course in the general population, data on all individuals aged 18 years and older who had received a ChAdOx1-S primary course and either a ChAdOx1-S or BNT162b2 booster dose were extracted from the NIMS on 14 March 2022.

To estimate vaccine effectiveness, testing data were linked to NIMS on 21 February 2021 using combinations of the unique individual National Health Service (NHS) number, date of birth, surname, first name, and postcode using deterministic linkage – 97.6% of eligible tests could be linked to the NIMS.

#### Identification of Delta and Omicron Variants and assignment to cases

Sequencing of PCR positive samples is undertaken through a network of laboratories, including the Wellcome Sanger Institute. Whole-genome sequences are assigned to UKHSA definitions of variants based on mutations (22, 23). S-gene target status on PCR-testing is an alternative approach for identifying variant because Delta has been associated with a positive S-gene target status on PCR testing with the Taqpath assay while Omicron BA.1 has been associated with S-gene target failure (SGTF). Omicron BA.2 has also been associated with positive S-gene target status but can be distinguished from Delta by sequencing and period. Cases were defined as Delta or Omicron (BA.1 and BA.2) based on whole genome sequencing, genotyping or SGTF, with sequencing taking priority, followed by genotyping and SGTF status. Where subsequent positive tests within 14 days included sequencing, genotyping or S-gene target failure information, this information was used to classify the variant. The Delta analysis was restricted from 13 September 2021 to 9 January 2022. The Omicron analysis was restricted from 29 November to 17 February 2022.

#### Hospital Admission Data

Hospital inpatient admissions for a range of acute respiratory illnesses were identified from the Secondary Uses Service (SUS) (24) and were linked to the testing data on 22 February 2022 using NHS number and date of birth. For the Pillar 2 samples, admissions with an ICD-10 acute respiratory illness (ARI) discharge diagnosis in the first diagnosis field (Supplementary Table 1) were identified where the sample was taken 14 days before and up to 2 days after the day of admission. For the Pillar 1 samples, admissions with an ICD-10 coded ARI discharge diagnosis in any diagnosis field were identified where the sample was taken 1 day before and up to 2 days after the admission. The data was restricted to tests up to 2 February 2022 to account for delays in the SUS data recording.

As a secondary analysis, Emergency Care hospital admissions from the Emergency Care Dataset (ECDS) (25), which includes hospital admissions through emergency departments but not elective admissions, were linked to the testing data using NHS number and date of birth on 22 February 2022 to identify admissions within 14 days of the sample date. The data were restricted to tests up to 6 February 2022 to allow sufficient follow-up time.

Since the milder Omicron variant became dominant, an increasing proportion of cases in hospital are likely to have COVID-19 as an incidental finding rather than as the primary reason for admission (26). Previously, we have found that using the ECDS data gives estimates that are likely more reflective of VE against infection (26). As such, we regard the SUS analysis as the primary analysis but also include the ECDS analysis to allow comparison to previous studies.

For a full description of the data sources, please see Supplementary Appendix.

### Statistical Analysis

Logistic regression was used to estimate differences in demographic and clinical characteristics of individuals who received a ChAdOx1-S booster as compared to those who received a BNT162b2 booster. The logistic regression model adjusted for age (ages 18-19, then 20 through to 89 in five-year bands, then everyone age 90 years or older), sex, index of multiple deprivation (quintile), ethnic group, geographic region (NHS region), health and social care worker status, clinical risk group status (only available for those aged 64 years and younger, clinically extremely vulnerable (CEV) status and severely immunosuppressed status (21, 27).

To estimate vaccine effectiveness, logistic regression was used with the PCR test result as the dependent variable and cases being those testing positive (stratified in separate analyses as either Omicron or Delta, and as either age 40 to 64 years or 65 years and older) and controls being those testing negative. Under 40-year olds were not included, given that ChAdOx1-S was not recommended in the general population in this age group. Vaccination status was included as an independent variable and effectiveness defined as 1-odds of vaccination in cases/odds of vaccination in controls. Vaccine effectiveness was adjusted in logistic regression models for age (ages 40 through to 89 in five-year bands, then everyone age 90 years or older), sex, index of multiple deprivation (quintile), ethnic group, history of travel, geographic region (NHS region), period (week of test), health and social care worker status, clinical risk group status, clinically extremely vulnerable, and previously testing positive. These factors were all considered potential confounders so were included in all models.

## Results

### Descriptive characteristics of those receiving ChAdOx1-S as a booster in England

To investigate the demographic characteristics of individuals who received a ChAdOx1-S booster vaccination, data were extracted from the NIMS on 14 March 2022 on all adults aged 18 years and older in England who had received a ChAdOx1-S primary course followed by a ChAdOx1-S or BNT162b2 booster (Table 1). In total, there were 43,171 individuals who received a ChAdOx1-S booster dose as compared to 13,038,908 individuals who received BNT162b2. Those who received a ChAdOx1-S booster were more likely to be female (adjusted odds ratio (OR) 1.67 (1.64-1.71)), from London (adjusted OR 2.84 (2.73-2.95), health and social care workers (adjusted OR 1.58 (1.54-1.63)), in a clinical risk group (adjusted OR 1.58 (1.54-1.63)), in the CEV group (adjusted OR 1.84 (1.79-1.89)) or in the severely immunosuppressed group (adjusted OR 2.05 (1.96-2.13)) (Table 1).

**Table 1.**
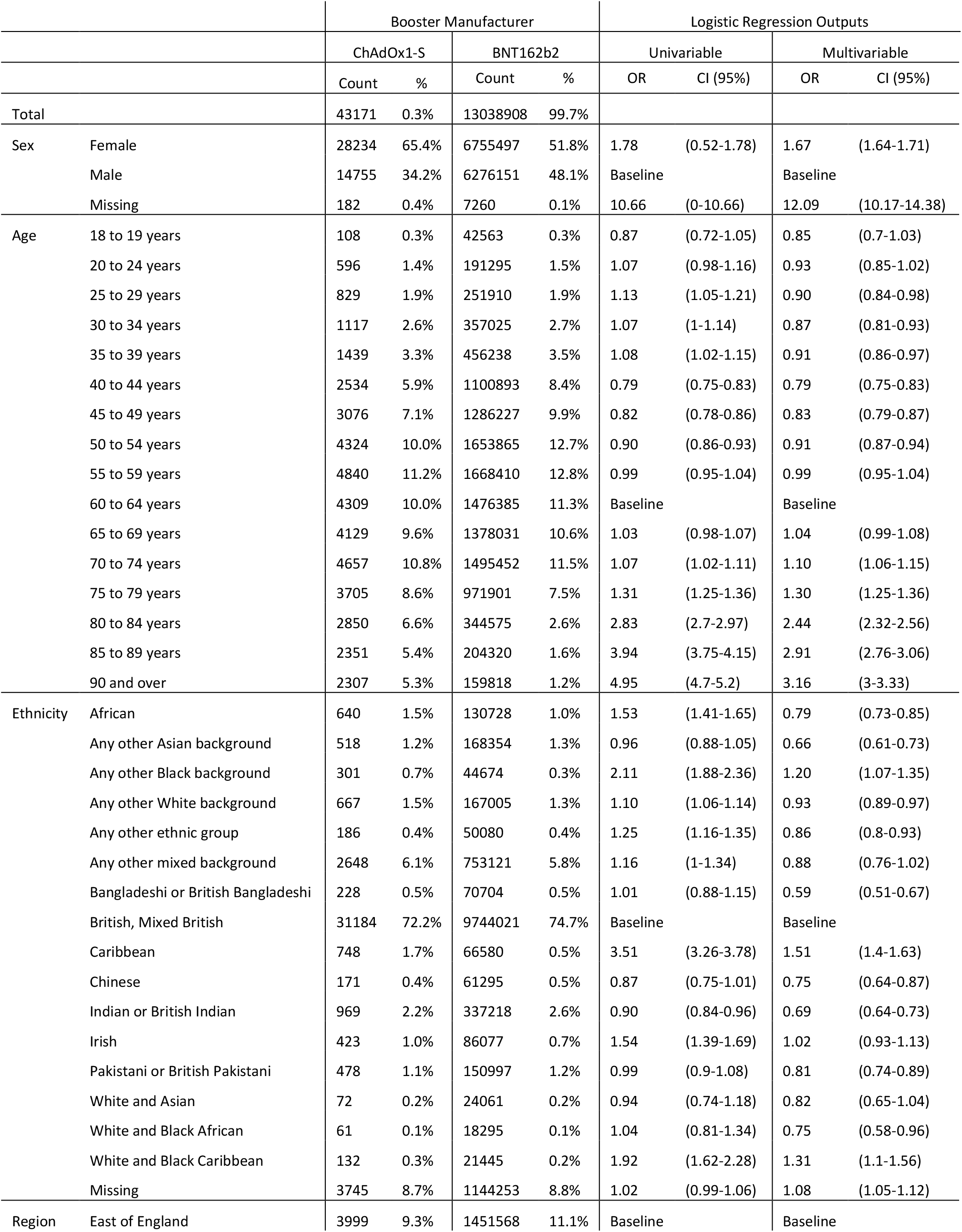
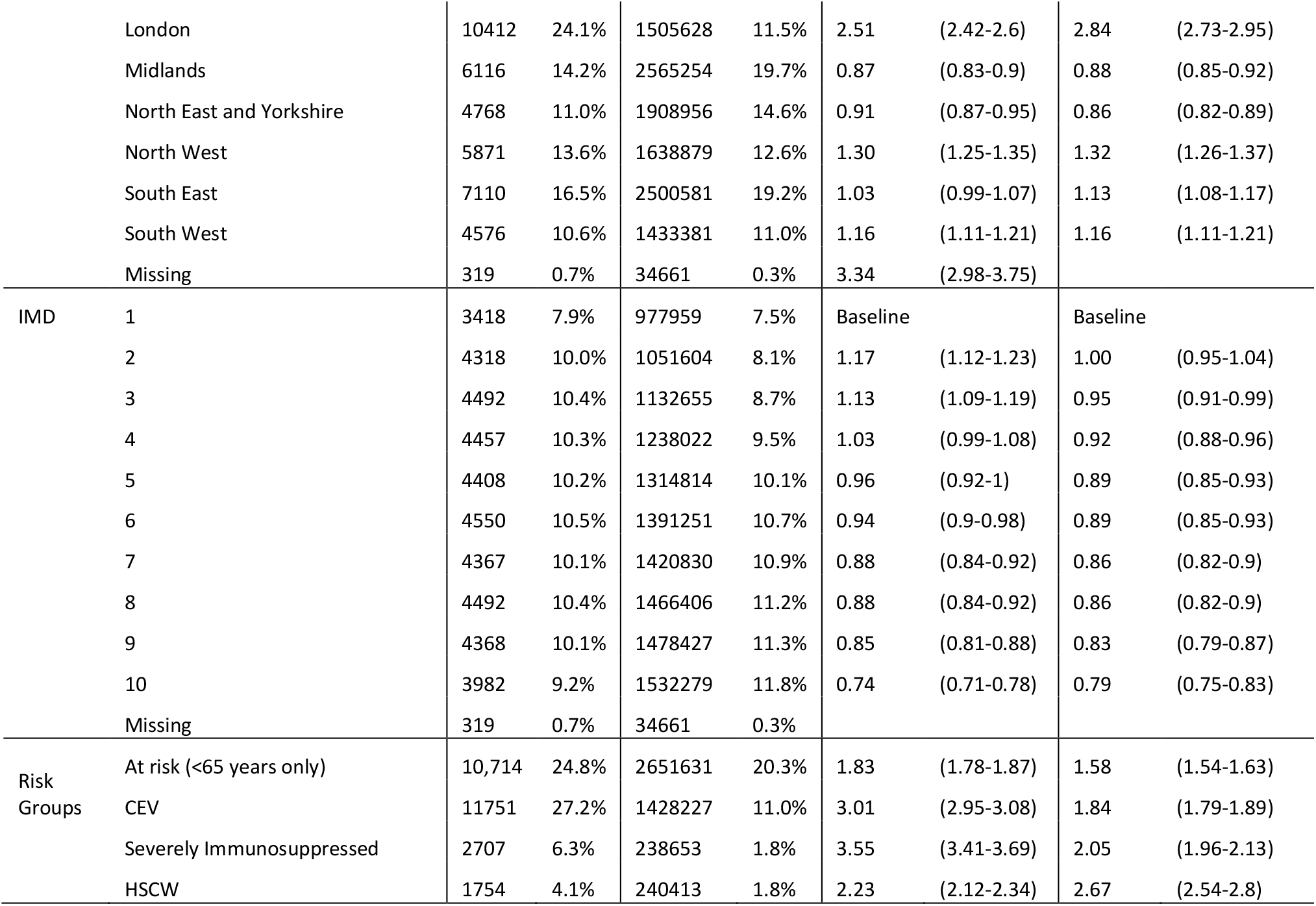
Demographic characteristics of the adults who received a ChAdOx1-S booster as compared to a BNT162b2 booster, following a ChAdOx1-S primary course, in England.

### Effectiveness of the ChAdOx1-S booster against symptomatic disease

Between the 29 November 2021 and 17 February 2022 (the Omicron variant study period), there was a total of 457,377 negative and 434,514 positive tests in those aged 40 years and older, with a test date within 10 days of the symptom onset date which could be linked to the National Immunisation Management system. A description of eligible tests from symptomatic cases is included in Supplementary Table 2.

Following a ChAdOx1-S primary vaccination course, VE against symptomatic disease caused by the Omicron variant was 8.0% (6.0 to 9.9%) and 19.5% (11.7 to 26.6%) for those aged 40-64 years and those aged 65 years and older, respectively, after 25 or more weeks (Table 2). In those aged 40-64 years, VE against symptomatic disease increased to 61.2% (40.9 to 74.6%) one week after receiving a ChAdOx1-S booster as compared to 58.2% (57 to 59.4%) amongst those who received a BNT162b2 booster. The protection of the ChAdOx1-S booster waned to 37.2% (−44.1 to 72.6%) at 15 or more weeks after receiving the booster, as compared to 30.6% (26.8 to 34.3%) over the same period amongst those who received a BNT162b2 booster.

**Table 2.** Effectiveness of the ChAdOx1-S and BNT162b2 booster vaccines against symptomatic disease following infection with the Omicron variant for adults aged 40 years and older in England.

| Age (years) | Dose | Booster Manufacturer | Interval (days) | Controls | Cases | OR* | VE (95% CI) |
| --- | --- | --- | --- | --- | --- | --- | --- |
| 40-64 | Unvaccinated |  |  | 27,361 | 51265 | Baseline | Baseline |
|  | Dose 2** | n/a | 175+ | 85175 | 89230 | 0.92 (0.9-0.94) | 8 (6 to 9.9) |
|  | Booster | Any*** | 0-1 | 11,879 | 7715 | 0.8 (0.77-0.83) | 20.3 (17.2 to 23.3) |
|  |  | Any*** | 2-6 | 27430 | 21422 | 0.74 (0.72-0.76) | 25.8 (23.7 to 27.8) |
|  |  | BNT162b2 | 7-13 | 28,809 | 17658 | 0.42 (0.41-0.43) | 58.2 (57.0 to 59.4) |
|  |  | BNT162b2 | 14-34 | 86719 | 66406 | 0.36 (0.35-0.37) | 63.8 (63.0 to 64.5) |
|  |  | BNT162b2 | 35-69 | 87592 | 90787 | 0.43 (0.42-0.44) | 57.3 (56.4 to 58.2) |
|  |  | BNT162b2 | 70-104 | 22504 | 29379 | 0.54 (0.52-0.55) | 46.4 (45.0 to 47.8) |
|  |  | BNT162b2 | 105+ | 2758 | 4278 | 0.69 (0.66-0.73) | 30.6 (26.8 to 34.3) |
|  |  | ChAdOx1-S | 7-13 | 70 | 40 | 0.39 (0.25-0.59) | 61.2 (40.9 to 74.6) |
|  |  | ChAdOx1-S | 14-34 | 193 | 159 | 0.48 (0.38-0.61) | 51.7 (38.9 to 61.8) |
|  |  | ChAdOx1-S | 35-69 | 216 | 215 | 0.47 (0.38-0.57) | 53.0 (42.6 to 61.6) |
|  |  | ChAdOx1-S | 70-104 | 69 | 97 | 0.59 (0.43-0.81) | 40.8 (18.6 to 56.9) |
|  |  | ChAdOx1-S | 105+ | 10 | 14 | 0.63 (0.27-1.44) | 37.2 (-44.1 to 72.6) |
| 65+ | Unvaccinated |  |  | 1,701 | 2361 | Baseline | Baseline |
|  | Dose 2** | n/a | 175+ | 4466 | 3053 | 0.81 (0.73-0.88) | 19.5 (11.7 to 26.6) |
|  | Booster | Any*** | 0-1 | 428 | 110 | 0.65 (0.5-0.85) | 34.6 (14.8 to 49.8) |
|  |  | Any*** | 2-6 | 1140 | 370 | 0.71 (0.61-0.84) | 28.6 (16.0 to 39.3) |
|  |  | BNT162b2 | 7-13 | 1,883 | 433 | 0.42 (0.36-0.48) | 58.1 (51.6 to 63.8) |
|  |  | BNT162b2 | 14-34 | 14311 | 3010 | 0.31 (0.29-0.34) | 68.5 (65.7 to 71.2) |
|  |  | BNT162b2 | 35-69 | 36300 | 25240 | 0.46 (0.42-0.49) | 54.1 (50.5 to 57.5) |
|  |  | BNT162b2 | 70-104 | 14210 | 18317 | 0.6 (0.55-0.65) | 40.1 (35.2 to 44.5) |
|  |  | BNT162b2 | 105+ | 1970 | 2789 | 0.77 (0.7-0.85) | 23.1 (15.1 to 30.5) |
|  |  | ChAdOx1-S | 7-13 | 23 | 8 | 0.34 (0.14-0.83) | 66.1 (16.6 to 86.3) |
|  |  | ChAdOx1-S | 14-34 | 53 | 32 | 0.48 (0.3-0.79) | 51.6 (20.8 to 70.4) |
|  |  | ChAdOx1-S | 35-69 | 88 | 81 | 0.56 (0.4-0.78) | 44.5 (22.4 to 60.2) |
|  |  | ChAdOx1-S | 70-104 | 16 | 40 | 1.27 (0.7-2.32) | -27.2 (-131.6 to 30.1) |
|  |  | ChAdOx1-S | 105+ | 3 | 5 | 0.98 (0.23-4.28) | N too low |

\*Odds ratio
\*\*ChAdOx1-S primary course
\*\*\*ChAdOx1-S or BNT162b2

Similar levels of protection against symptomatic disease was observed in those aged 65 years and older (Table 2). After receiving the booster, VE peaked at 66.1% (16.6 to 86.3%) and 68.5% (65.7 to 71.2%) amongst those who received the ChAdOx1-S and BNT162b2 booster vaccines, respectively. In older adults, this protection waned to 44.5% (22.4 to 60.2%) and 54.1% (50.5 to 57.5%) after 5-9 weeks amongst those who received the ChAdOx1-S and BNT162b2 booster vaccines, respectively. Beyond 10 weeks the analysis was limited by small numbers but there was evidence of further waning.

### Effectiveness of the ChAdOx1-S booster against hospitalisation

The effectiveness of the ChAdOx1-S booster against hospitalisation was estimated following infection with either Delta or Omicron variants, by linkage of Pillar 1 and Pillar 2 tests to the secondary care (SUS) data (Table 3, Figure 1) and by linkage of Pillar 2 tests to accident and emergency (ECDS) data (Supplementary Table 3).

**Table 3.**
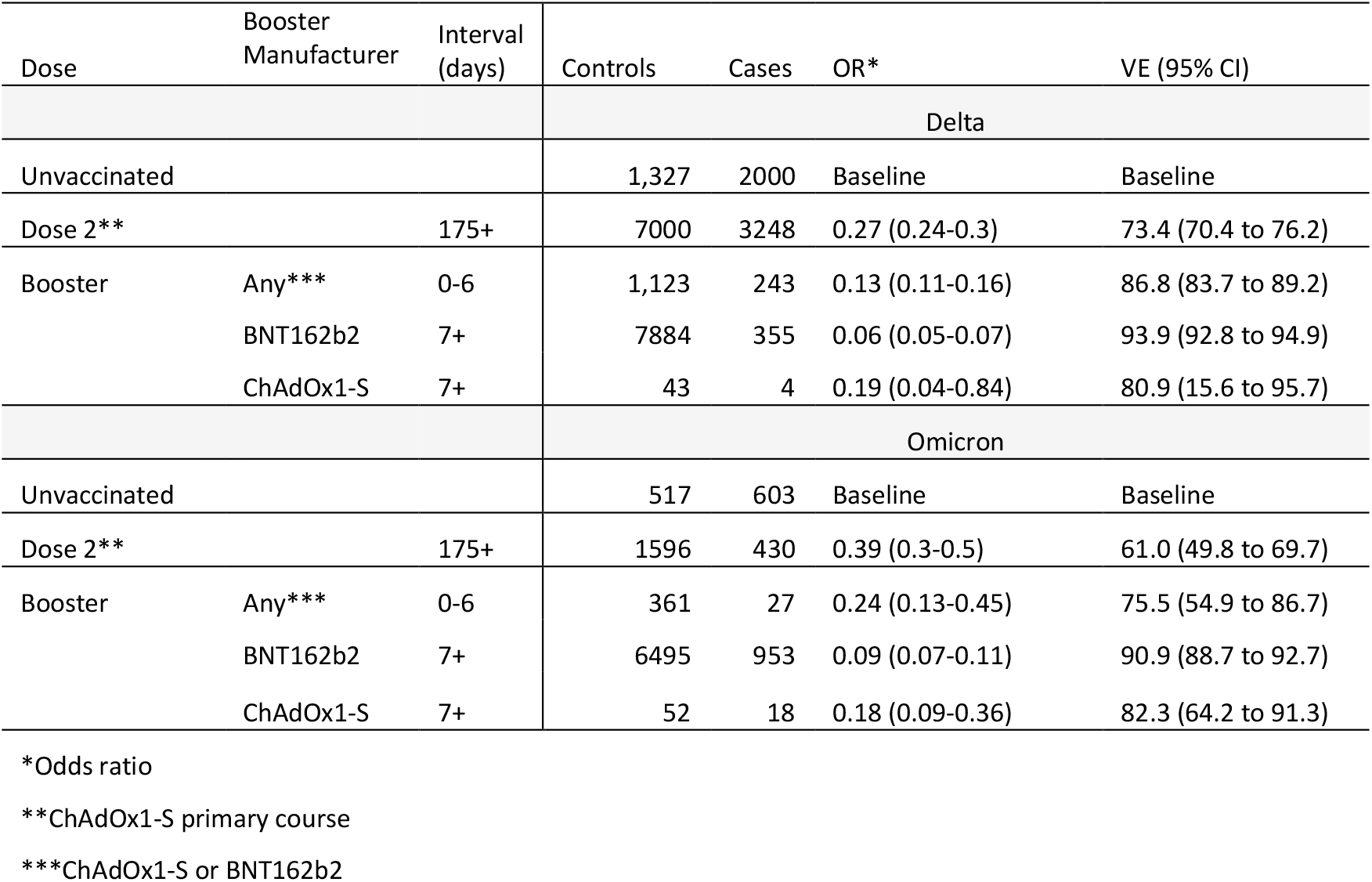
Effectiveness of the ChAdOx1-S and BNT162b2 booster vaccines against hospitalisation as defined by linkage to the secondary care (SUS) data following infection with Delta or Omicron variants in adults aged 65 years and older in England.

| Dose | Booster Manufacturer | Interval (days) | Controls | Cases | OR* | VE (95% CI) |
| --- | --- | --- | --- | --- | --- | --- |
| Delta |  |  |  |  |  |  |
| Unvaccinated |  |  | 1,327 | 2000 | Baseline | Baseline |
| Dose 2** |  | 175+ | 7000 | 3248 | 0.27 (0.24-0.3) | 73.4 (70.4 to 76.2) |
| Booster | Any*** | 0-6 | 1,123 | 243 | 0.13 (0.11-0.16) | 86.8 (83.7 to 89.2) |
|  | BNT162b2 | 7+ | 7884 | 355 | 0.06 (0.05-0.07) | 93.9 (92.8 to 94.9) |
|  | ChAdOx1-S | 7+ | 43 | 4 | 0.19 (0.04-0.84) | 80.9 (15.6 to 95.7) |
| Omicron |  |  |  |  |  |  |
| Unvaccinated |  |  | 517 | 603 | Baseline | Baseline |
| Dose 2** |  | 175+ | 1596 | 430 | 0.39 (0.3-0.5) | 61.0 (49.8 to 69.7) |
| Booster | Any*** | 0-6 | 361 | 27 | 0.24 (0.13-0.45) | 75.5 (54.9 to 86.7) |
|  | BNT162b2 | 7+ | 6495 | 953 | 0.09 (0.07-0.11) | 90.9 (88.7 to 92.7) |
|  | ChAdOx1-S | 7+ | 52 | 18 | 0.18 (0.09-0.36) | 82.3 (64.2 to 91.3) |

\*Odds ratio
\*\*ChAdOx1-S primary course
\*\*\*ChAdOx1-S or BNT162b2

**Figure 1.**
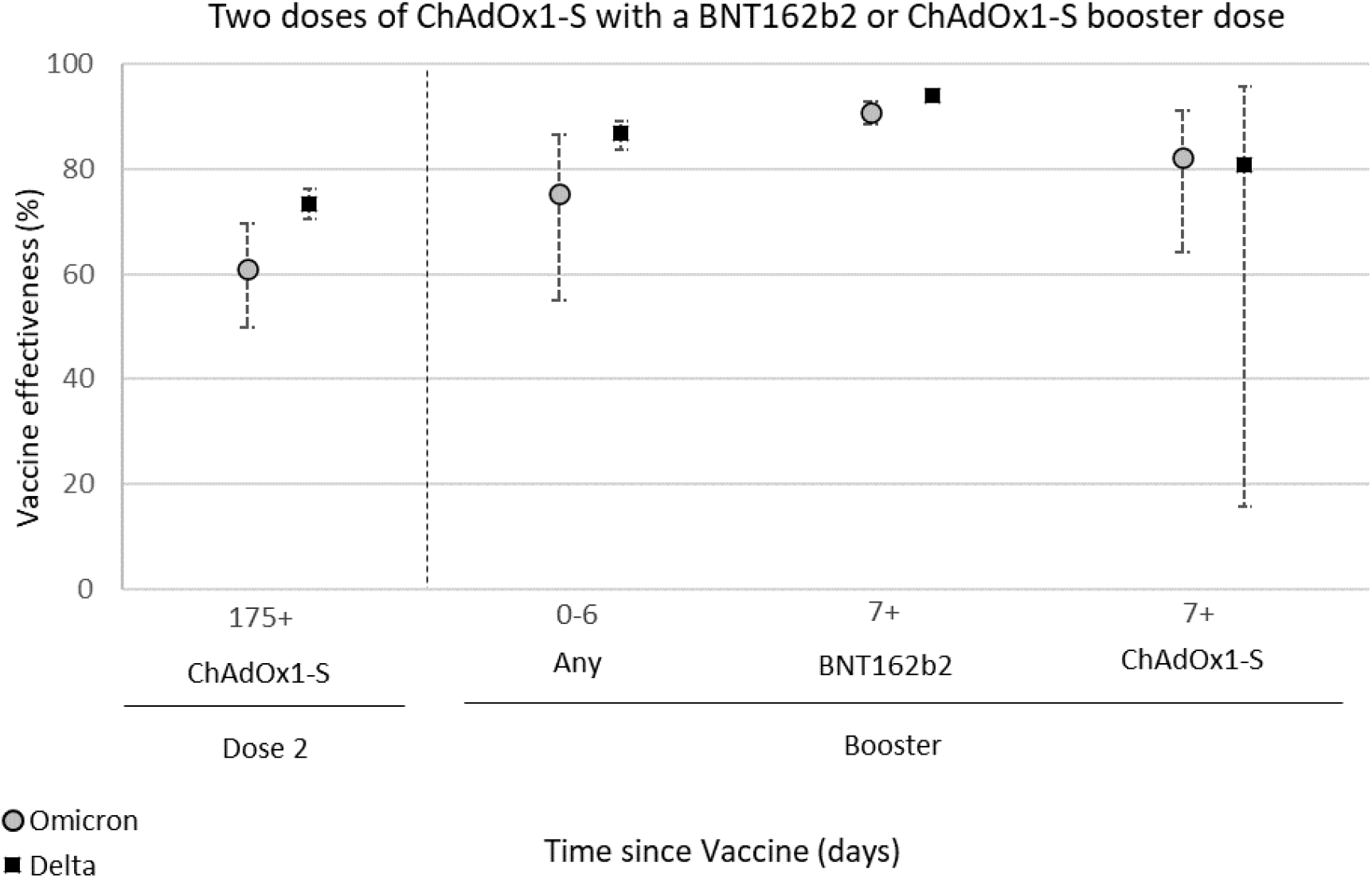
Effectiveness of the ChAdOx1-S and BNT162b2 booster vaccines against hospitalisation as defined by linkage to the secondary care (SUS) data following infection with Delta or Omicron variants in those aged 65 years and older in England.

Following linkage to the SUS data, there were a total of 17,377 negative and 5,850 positive (Delta) eligible tests between 13 September 2021 and 9 January 2022, while there were a total of 9,021 negative and 2,031 positive (Omicron) eligible tests between 29 November 2021 and 2 February 2022. Following linkage to the ECDS data, there was a total of 469,976 negative and 1,352 positive (Delta) eligible tests between 13 September 2021 and 9 January 2022, while there was a total of 457,377 negative and 509 positive (Omicron) eligible tests between 29 November 2021 and 17 February 2022. A description of eligible tests from all hospitalised cases is included in Supplementary Tables 2&4.

Amongst those aged 65 years and older, protection against hospitalisation (defined as requiring a stay of two or more days with severe respiratory disease) following Omicron infection was 61.0% (49.8 to 69.7%) at 25 weeks after a second dose (Table 3, Figure 1). This increased to 82.3% (64.2 to 91.3%) one or more weeks after receiving a ChAdOx1-S booster, as compared to 90.9% (88.7 to 92.7%) for those who received a BNT162b2 booster. Protection against hospitalisation following Delta infection was also enhanced by the ChAdOx1-S booster in older adults. VE against Delta hospitalisation was 73.4% (70.4 to 76.2%) at 25 weeks after receiving a second dose. This increased to 80.9% (15.6% to 95.7%) and 93.9% (92.8% to 94.9%) respectively one week after vaccination with the ChAdOx1-S and BNT162b2 booster vaccines, respectively (Table 3, Figure 1). VE estimates derived using a broader definition of hospitalisation with the ECDS-linked data after a booster dose of ChAdOx1-S were substantially lower at around 65% with either variant (Supplementary Table 3).

## Discussion

Here, we find reassuring real-world evidence that booster vaccination with ChAdOx1-S following a primary ChAdOx1-S course provides increased protection against mild and severe disease with Delta and Omicron infection. Protection against symptomatic disease with Omicron was broadly similar to that seen with BNT162b2, with substantial waning by 10 or more weeks. Vaccine effectiveness against hospitalisation was high after a booster dose of ChAdOx1-S at over 80%, though lower than that observed following a BNT162b2 booster.

Boosting with ChAdOx1-S is not recommended as standard in the UK COVID-19 vaccination programme and there were notable differences between individuals who received ChAdOx1-S and BNT162b2 booster doses. Those who received three doses of ChAdOx1-S were more likely to be in risk groups; this is in line with the national recommendation that only those who were clinically contraindicated to get an mRNA booster should get a ChAdOx1-S booster (21). From previous studies in the UK, we know that VE is lower in the clinical vulnerable populations (14). Although this risk status was adjusted for in the VE estimates, there is likely to be residual confounding which could have lowered the estimates of the ChAdOx1-S booster. Direct comparison of the protection conferred by the ChAdOx1-S as compared to BNT162b2 vaccination is therefore very challenging using data from England. Furthermore, very few people received a ChAdOx1-S booster as compared to a BNT162b2 booster, and the confidence intervals around the estimates for this group were large.

Nonetheless, boosting with ChAdOx1-S improved protection against symptomatic disease following Omicron infection from the waning observed at 25 weeks post a second dose in both younger (40 to 64 years) and older (65 years and older) adults. Our findings are consistent with data from COV-BOOST in the UK and from Thailand which found both homologous and heterologous boosting with ChAdOx1-S enhanced immunity as measured by anti-spike IgG and neutralisation assays (6, 16). Although booster vaccination with both BNT162b2 and ChAdOx1-S enhanced immunity, data from COV-BOOST found that boosting with BNT162b2 resulted in higher titres of anti-spike IgG, a greater cellular response as well as increased neutralising capacity as compared to ChAdOx1-S (16). Early real-world data from Brazil also indicate that the ChAdOx1-S booster confers a high level of protection against severe disease (28). Together, these data support the use of the ChAdOx1-S booster vaccines to prevent severe disease, including against the Omicron variant. Significant waning was observed in the period after booster vaccination, indicating that further boosters will be required if the aim of a vaccination programme is to prevent infection or mild disease.

The Omicron variant has previously been demonstrated to evade immunity from previous infection and vaccination (1), however reassuringly the ChAdOx1-S booster still provided high levels of protection against severe disease with this variant. Previously we have found that using broader definitions of hospitalisation has given lower VE estimates, reflecting outcome misclassification where cases are likely coincidentally positive whilst in hospital, without this being the reason for admission (26). This is likely to be the reason for the lower estimates using the ECDS data. Protection against severe disease defined more specifically as requiring at least two days stay in hospital and a respiratory code in the primary diagnostic field using the SUS data demonstrated that protection from severe disease was very high amongst those who received the ChAdOx1-S booster. Numbers were too small to look at more severe and specific endpoints such as those who required oxygen, ventilation, or intensive care. In previous studies, using these endpoints yielded higher vaccine effectiveness estimates (26).

This study supports the use of ChAdOx1-S as a booster for protection against severe disease with COVID-19 in settings that have not yet offered booster doses and suggests that those who received ChAdOx1-S as a booster in the UK do not require re-vaccination ahead of others. Comparison with other vaccines is challenging due to the different populations that have received each vaccine in the UK and head to head trials are likely to be required for a robust comparison.

## Data Availability

All data produced in the present work are contained in the manuscript.

## Authors’ contributions

FCMK and JLB wrote the manuscript. JLB, NA conceptualised the study. FCMK, JS, RS curated the data. RS, NA, FCMK conducted the demographic analysis. NA conducted the VE analysis. All co-authors reviewed the manuscript.

## Ethics committee approval

UKHSA Research Ethics and Governance Group Statement: Surveillance of COVID-19 testing and vaccination is undertaken under Regulation 3 of The Health Service (Control of Patient Information) Regulations 2002 to collect confidential patient information (http://www.legislation.gov.uk/uksi/2002/1438/regulation/3/made) under Sections 3(i) (a) to (c), 3(i)(d) (i) and (ii) and 3(3). The study protocol was subject to an internal review by the PHE Research Ethics and Governance Group and was found to be fully compliant with all regulatory requirements. As no regulatory issues were identified, and ethical review is not a requirement for this type of work, it was decided that a full ethical review would not be necessary.

All necessary patient/participant consent has been obtained and the appropriate institutional forms have been archived.

## References

1. Andrews N, Stowe J, Kirsebom F, Toffa S, Rickeard T, Gallagher E, et al. Covid-19 Vaccine Effectiveness against the Omicron (B.1.1.529) Variant. New England Journal of Medicine. 2022.

2. Andrews N, Stowe J, Kirsebom F, Toffa S, Sachdeva R, Gower C, et al. Effectiveness of COVID-19 booster vaccines against covid-19 related symptoms, hospitalisation and death in England. Nature medicine. 2022.

3. Bar-On YM, Goldberg Y, Mandel M, Bodenheimer O, Freedman L, Kalkstein N, et al. Protection of BNT162b2 Vaccine Booster against Covid-19 in Israel. New England Journal of Medicine. 2021;385(15):1393–400.

4. Arbel R, Hammerman A, Sergienko R, Friger M, Peretz A, Netzer D, et al. BNT162b2 Vaccine Booster and Mortality Due to Covid-19. New England Journal of Medicine. 2021;385(26):2413–20.

5. Our World in Data: COVID-19 vaccine boosters administered [Available from: https://ourworldindata.org/covid-vaccinations.

6. Kanokudom S, Assawakosri S, Suntronwong N, Auphimai C, Nilyanimit P, Vichaiwattana P, et al. Safety and Immunogenicity of the Third Booster Dose with Inactivated, Viral Vector, and mRNA COVID-19 Vaccines in Fully Immunized Healthy Adults with Inactivated Vaccine. Vaccines (Basel). 2022;10(1):86.

7. Hart JD, Chokephaibulkit K, Mayxay M, Ong-Lim Alt, Saketa ST, Russell FM. COVID-19 vaccine boosters in the Asia-Pacific region in the context of Omicron. The Lancet Regional Health – Western Pacific. 2022;20.

8. VIEW-hub by IVAC [Available from: https://view-hub.org/covid-19/.

9. Andrews N, Stowe J, Kirsebom F, Toffa S, Rickeard T, Gallagher E, et al. Effectiveness of COVID-19 vaccines against the Omicron (B.1.1.529) variant of concern. medRxiv. 2021:2021.12.14.21267615.

10. Lopez Bernal J, Andrews N, Gower C, Gallagher E, Simmons R, Thelwall S, et al. Effectiveness of Covid-19 Vaccines against the B.1.617.2 (Delta) Variant. New England Journal of Medicine. 2021;385(7):585–94.

11. Thiruvengadam R, Awasthi A, Medigeshi G, Bhattacharya S, Mani S, Sivasubbu S, et al. Effectiveness of ChAdOx1 nCoV-19 vaccine against SARS-CoV-2 infection during the delta (B.1.617.2) variant surge in India: a test-negative, case-control study and a mechanistic study of post-vaccination immune responses. (1474–4457 (Electronic)).

12. Madhi SA, Baillie V, Cutland CL, Voysey M, Koen AL, Fairlie L, et al. Efficacy of the ChAdOx1 nCoV-19 Covid-19 Vaccine against the B.1.351 Variant. New England Journal of Medicine. 2021;384(20):1885–98.

13. Voysey M, Clemens SAC, Madhi SA, Weckx LY, Folegatti PM, Aley PK, et al. Safety and efficacy of the ChAdOx1 nCoV-19 vaccine (AZD1222) against SARS-CoV-2: an interim analysis of four randomised controlled trials in Brazil, South Africa, and the UK. The Lancet. 2020.

14. Andrews N, Tessier E, Stowe J, Gower C, Kirsebom F, Simmons R, et al. Duration of Protection against Mild and Severe Disease by Covid-19 Vaccines. New England Journal of Medicine. 2022.

15. UK Health Security Agency. COVID-19 vaccine surveillance report: Week 11 2022 [Available from: https://assets.publishing.service.gov.uk/government/uploads/system/uploads/attachment_data/file/1061532/Vaccine_surveillance_report_-_week_11.pdf.

16. Munro APS, Janani L, Cornelius V, Aley PK, Babbage G, Baxter D, et al. Safety and immunogenicity of seven COVID-19 vaccines as a third dose (booster) following two doses of ChAdOx1 nCov-19 or BNT162b2 in the UK (COV-BOOST): a blinded, multicentre, randomised, controlled, phase 2 trial. The Lancet. 2021;398(10318):2258–76.

17. Flaxman A, Marchevsky NG, Jenkin D, Aboagye J, Aley PK, Angus B, et al. Reactogenicity and immunogenicity after a late second dose or a third dose of ChAdOx1 nCoV-19 in the UK: a substudy of two randomised controlled trials (COV001 and COV002). The Lancet. 2021;398(10304):981–90.

18. Costa Clemens SA, Weckx L, Clemens R, Almeida Mendes AV, Ramos Souza A, Silveira MBV, et al. Heterologous versus homologous COVID-19 booster vaccination in previous recipients of two doses of CoronaVac COVID-19 vaccine in Brazil (RHH-001): a phase 4, non-inferiority, single blind, randomised study. The Lancet. 2022;399(10324):521–9.

19. Greinacher A, Thiele T, Warkentin TE, Weisser K, Kyrle PA, Eichinger S. Thrombotic Thrombocytopenia after ChAdOx1 nCov-19 Vaccination. New England Journal of Medicine. 2021;384(22):2092–101.

20. Joint Committee on Vaccination and Immunisation. JCVI advises on COVID-19 vaccine for people aged under 40 2021 [Available from: https://www.gov.uk/government/news/jcvi-advises-on-covid-19-vaccine-for-people-aged-under-40.

21. UK Health Security Agency. COVID-19: the green book, chapter 14a. Immunisation against infectious diseases: UK Health Security Agency,; 2020.

22. UK Health Security Agency. SARS-CoV-2 variants of concern and variants under investigation in England Variant of concern: Omicron, VOC21NOV-01 (B.1.1.529) Technical briefing 30 2021 [Available from: https://assets.publishing.service.gov.uk/government/uploads/system/uploads/attachment_data/file/1038404/Technical_Briefing_30.pdf.

23. Bull M, Chand M, Connor T, Ellaby N, Groves N, Jalava K, et al. Standardised Variant Definitions 2022 [Available from: https://github.com/phe-genomics/variant_definitions.

24. NHS Digital. Secondary Uses Service (SUS) 2022 [Available from: https://digital.nhs.uk/services/secondary-uses-service-sus.

25. NHS Digital. Emergency Care Data Set (ECDS) 2022 [Available from: https://digital.nhs.uk/data-and-information/data-collections-and-data-sets/data-sets/emergency-care-data-set-ecds.

26. Stowe J, Andrews N, Kirsebom F, Ramsay M, Bernal JL. Effectiveness of COVID-19 vaccines against Omicron and Delta hospitalisation: test negative case-control study. medRxiv. 2022:2022.04.01.22273281.

27. NHS Digital. COVID-19 – high risk shielded patient list identification methodology 2020 [Available from: https://digital.nhs.uk/coronavirus/shielded-patient-list/methodology/rule-logic.

28. Cerqueira-Silva T, de Araujo Oliveira V, Paixão ES, Veras Florentino PT, Penna GO, Pearce N, et al. Protection conferred by vaccine plus previous infection (hybrid immunity) with vaccines of three different platforms during the Omicron variant period in Brazil. medRxiv. 2022:2022.04.12.22273752.

