## Supplementary Appendix for "Effectiveness of ChAdOx1-S COVID-19 Booster Vaccination against the Omicron and Delta variants in England"

### Supplementary Methods

#### Data Sources

##### COVID-19 Testing Data
SARS-CoV-2 Testing PCR testing for SARS CoV-2 in England is undertaken by hospital and public health laboratories (Pillar 1), as well as by community testing (Pillar 2). Pillar 1 testing data is available from UKHSA labs and NHS hospitals for those with a clinical need, and health and care workers. Pillar 2 community testing is available to anyone with symptoms consistent with COVID-19 (high temperature, new continuous cough, or loss or change in sense of smell or taste), anyone who is a contact of a confirmed case, care home staff and residents, and to those who have self-tested as positive using a lateral flow test (LFT).

##### Vaccination Data
The National Immunization Management System (NIMS) contains demographic information on the whole population of England who are registered with a general practice physician in England and is used to record all COVID-19 vaccinations. NIMS was accessed for dates of vaccination and manufacturer, sex, date of birth, ethnicity, and residential address. Addresses were used to determine index of multiple deprivation quintile and were also linked to Care Quality Commission registered care homes using the unique property reference number. Data on geography (NHS region), risk group status, clinically extremely vulnerable status, and health/social care worker were also extracted from the NIMS. Clinical risk groups included a range of chronic conditions as described in the Green Book (1), whereas the clinically extremely vulnerable group included persons who were considered to be at the highest risk for severe COVID-19, including those with immunosuppressed conditions and those with severe respiratory disease. Booster doses were identified as a third dose given at least 84 days after a second dose and administered after 13 September 2021. Individuals with four or more doses of vaccine, heterologous primary schedule or fewer than 19 days between their first and second dose were excluded.

##### Emergency Care Hospital Admission Data
Admissions due to an injury were excluded. Admissions were identified where the Emergency Care Destination code was either discharge to a ward, intensive care unit, coronary care unit, high dependency unit or where there was a date on which the decision to admit the patient was made. Admissions with an acute respiratory illness (ARI) SNOMED coded as the reason for attending emergency care were flagged.

##### Secondary Care Hospital Admission Data
SUS is the national electronic database of hospital admissions that provides timely updates of ICD-10 codes for completed hospital stays for all NHS hospitals in England. Up to 24 ICD-10 diagnoses fields can be completed in SUS for each admission with the first diagnosis field indicating the primary reason for admission. Length of stay was calculated as date of discharge – date of admission. Where multiple admissions linked to the same sample date the first admission after the sample date was retained and episode length calculated by summing the stay length for each admission. Data were

restricted to those with ARI in the first diagnosis field and where the length of stay was at least two days.

### Control selection

For analyses involving hospitalised controls any negative tests that led to a hospitalisation within 21 days of a previous hospital negative test were excluded. A maximum of one negative test per person within each of the following approximate 3-month periods was selected at random: 26 April to 1 August 2021, 2 August 2021 to 21 November 2021, 22 November 2021 to 2 February 2022. For analyses involving all Pillar 2 symptomatic controls the same was done within this control group.

### Supplementary Tables

Supplementary Table 1. SUS Acute respiratory illness ICD10 code list.

| SUS Acute respiratory illness ICD10 code list | |
|---|---|
| J04* | Acute laryngitis and tracheitis |
| J09* | Influenza due to identified avian influenza virus |
| J10* | Influenza with pneumonia, other influenza virus identified |
| J11* | Influenza with pneumonia, virus not identified |
| J12* | Viral pneumonia, not elsewhere classified |
| J13* | Pneumonia due to Streptococcus pneumoniae |
| J14* | Pneumonia due to Haemophilus influenzae |
| J15* | Bacterial pneumonia, not elsewhere classified |
| J16* | Pneumonia due to other infectious organisms, not elsewhere classified |
| J17* | Pneumonia in diseases classified elsewhere |
| J18* | Pneumonia, organism unspecified |
| J20* | Acute bronchitis |
| J21* | Acute bronchiolitis |
| J22* | Unspecified acute lower respiratory infection |
| J80* | ARDS (related to respiratory infection) |
| U07* | COVID-19, virus identified and not identified |
| U04* | Severe acute respiratory syndrome (SARS) |

Supplementary Table 2. Descriptive characteristics of those included in the VE analysis for protection against symptomatic disease (cases all) and hospitalisation as assessed using the ECDS data following infection with either Delta or Omicron variants.

| | | | | Delta | | | | | | Omicron | | | | | |
|---|---|---|---|---|---|---|---|---|---|---|---|---|---|---|---|
| | | | | Controls | | Cases All | | Cases Hosp | | Controls | | Cases All | | Cases Hosp | |
| | Dose | Booster manufacturer | Interval (days) | n | % | n | % | n | % | n | % | n | % | n | % |
| | | | | 469,976 | | 172,223 | | 1,352 | | 457,377 | | 434,514 | | 509 | |
| Vaccination Status | Unvaccinated | | | 49,918 | 10.6% | 48,699 | 28.3% | 366 | 27.1% | 29,062 | 6.4% | 53,626 | 12.3% | 93 | 18.3% |
| | Dose 2* | | 175+ | 142,454 | 30.3% | 92,917 | 54.0% | 809 | 59.8% | 89,641 | 19.6% | 92,283 | 21.2% | 84 | 16.5% |
| | Booster | Any | 0-6 | 51,109 | 10.9% | 21,257 | 12.3% | 96 | 7.1% | 40,877 | 8.9% | 29,617 | 6.8% | 5 | 1.0% |
| | Booster | BNT162b2 | 7+ | 225,962 | 48.1% | 9,303 | 5.4% | 76 | 5.6% | 297,056 | 64.9% | 258,297 | 59.4% | 320 | 62.9% |
| | Booster | ChAdOx1-S | 7+ | 533 | 0.1% | 47 | 0.0% | 5 | 0.4% | 741 | 0.2% | 691 | 0.2% | 7 | 1.4% |
| Gender | Female | | | 284,136 | 60.5% | 91,669 | 53.2% | 624 | 46.2% | 274,565 | 60.0% | 230,789 | 53.1% | 250 | 49.1% |
| | Male | | | 185,025 | 39.4% | 80,291 | 46.6% | 728 | 53.8% | 182,025 | 39.8% | 203,031 | 46.7% | 258 | 50.7% |
| | Missing | | | 815 | 0.2% | 263 | 0.2% | 0 | 0.0% | 787 | 0.2% | 694 | 0.2% | 1 | 0.2% |
| Age | 40-44 | | | 80,921 | 17.2% | 35,639 | 20.7% | 0 | 0.0% | 83,475 | 18.3% | 94,518 | 21.8% | 0 | 0.0% |
| | 45-49 | | | 74,782 | 15.9% | 30,300 | 17.6% | 0 | 0.0% | 78,924 | 17.3% | 86,768 | 20.0% | 0 | 0.0% |
| | 50-54 | | | 89,098 | 19.0% | 32,587 | 18.9% | 0 | 0.0% | 90,779 | 19.8% | 87,744 | 20.2% | 0 | 0.0% |
| | 55-59 | | | 76,254 | 16.2% | 25,591 | 14.9% | 0 | 0.0% | 75,520 | 16.5% | 67,858 | 15.6% | 0 | 0.0% |
| | 60-64 | | | 55,023 | 11.7% | 18,038 | 10.5% | 0 | 0.0% | 52,087 | 11.4% | 41,777 | 9.6% | 0 | 0.0% |
| | 65-69 | | | 40,628 | 8.6% | 12,894 | 7.5% | 405 | 30.0% | 34,908 | 7.6% | 24,659 | 5.7% | 116 | 22.8% |
| | 70-74 | | | 30,900 | 6.6% | 10,104 | 5.9% | 405 | 30.0% | 24,787 | 5.4% | 18,125 | 4.2% | 134 | 26.3% |
| | 75-79 | | | 14,709 | 3.1% | 4,876 | 2.8% | 275 | 20.3% | 11,152 | 2.4% | 8,756 | 2.0% | 103 | 20.2% |
| | 80-84 | | | 4,094 | 0.9% | 1,269 | 0.7% | 144 | 10.7% | 3,064 | 0.7% | 2,332 | 0.5% | 64 | 12.6% |
| | 85-89 | | | 1,999 | 0.4% | 580 | 0.3% | 81 | 6.0% | 1,518 | 0.3% | 1,107 | 0.3% | 49 | 9.6% |
| | 90+ | | | 1,568 | 0.3% | 345 | 0.2% | 42 | 3.1% | 1,163 | 0.3% | 870 | 0.2% | 43 | 8.4% |
| Ethnicity | African | | | 3,922 | 0.8% | 1,291 | 0.7% | 14 | 1.0% | 3,701 | 0.8% | 5,492 | 1.3% | 5 | 1.0% |
| | Any other Asian background | | | 5,405 | 1.2% | 1,668 | 1.0% | 14 | 1.0% | 5,437 | 1.2% | 6,253 | 1.4% | 6 | 1.2% |
| | Any other Black background | | | 1,810 | 0.4% | 787 | 0.5% | 4 | 0.3% | 1,590 | 0.3% | 2,617 | 0.6% | 0 | 0.0% |

| | | | | | | | | | | | | | |
|---|---|---|---|---|---|---|---|---|---|---|---|---|---|
| | Any other White background | 30,310 | 6.4% | 16,093 | 9.3% | 89 | 6.6% | 29,473 | 6.4% | 39,506 | 9.1% | 29 | 5.7% |
| | Any other ethnic group | 6,239 | 1.3% | 2,579 | 1.5% | 12 | 0.9% | 6,242 | 1.4% | 7,630 | 1.8% | 10 | 2.0% |
| | Any other mixed background | 1,945 | 0.4% | 774 | 0.4% | 7 | 0.5% | 1,894 | 0.4% | 2,038 | 0.5% | 2 | 0.4% |
| | Bangladeshi or British Bangladeshi | 1,743 | 0.4% | 678 | 0.4% | 9 | 0.7% | 1,741 | 0.4% | 2,198 | 0.5% | 4 | 0.8% |
| | British, Mixed British | 355,004 | 75.5% | 123,459 | 71.7% | 1,036 | 76.6% | 345,349 | 75.5% | 302,173 | 69.5% | 383 | 75.2% |
| | Caribbean | 2,847 | 0.6% | 1,443 | 0.8% | 16 | 1.2% | 2,474 | 0.5% | 4,463 | 1.0% | 9 | 1.8% |
| | Chinese | 1,451 | 0.3% | 490 | 0.3% | 7 | 0.5% | 1,503 | 0.3% | 1,453 | 0.3% | 3 | 0.6% |
| | Indian or British Indian | 12,668 | 2.7% | 3,191 | 1.9% | 29 | 2.1% | 12,322 | 2.7% | 11,073 | 2.5% | 7 | 1.4% |
| | Irish | 3,313 | 0.7% | 921 | 0.5% | 7 | 0.5% | 3,170 | 0.7% | 2,649 | 0.6% | 10 | 2.0% |
| | Pakistani or British Pakistani | 5,791 | 1.2% | 2,145 | 1.2% | 19 | 1.4% | 5,770 | 1.3% | 7,135 | 1.6% | 10 | 2.0% |
| | White and Asian | 832 | 0.2% | 301 | 0.2% | 0 | 0.0% | 865 | 0.2% | 834 | 0.2% | 0 | 0.0% |
| | White and Black African | 656 | 0.1% | 255 | 0.1% | 0 | 0.0% | 655 | 0.1% | 850 | 0.2% | 1 | 0.2% |
| | White and Black Caribbean | 915 | 0.2% | 404 | 0.2% | 4 | 0.3% | 840 | 0.2% | 1,284 | 0.3% | 2 | 0.4% |
| | Missing | 35,125 | 7.5% | 15,744 | 9.1% | 85 | 6.3% | 34,351 | 7.5% | 36,866 | 8.5% | 28 | 5.5% |
| NHS Region | East of England | 55,371 | 11.8% | 19,965 | 11.6% | 127 | 9.4% | 54,106 | 11.8% | 44,943 | 10.3% | 46 | 9.0% |
| | London | 56,857 | 12.1% | 18,455 | 10.7% | 121 | 8.9% | 55,327 | 12.1% | 61,545 | 14.2% | 59 | 11.6% |
| | Midlands | 89,571 | 19.1% | 33,599 | 19.5% | 274 | 20.3% | 87,072 | 19.0% | 82,976 | 19.1% | 100 | 19.6% |
| | North East | 70,243 | 14.9% | 27,797 | 16.1% | 287 | 21.2% | 66,238 | 14.5% | 69,399 | 16.0% | 98 | 19.3% |
| | North West | 65,199 | 13.9% | 23,730 | 13.8% | 180 | 13.3% | 60,400 | 13.2% | 71,916 | 16.6% | 101 | 19.8% |
| | South East | 82,624 | 17.6% | 28,623 | 16.6% | 210 | 15.5% | 85,855 | 18.8% | 70,342 | 16.2% | 69 | 13.6% |
| | South West | 50,110 | 10.7% | 20,054 | 11.6% | 153 | 11.3% | 48,378 | 10.6% | 33,393 | 7.7% | 36 | 7.1% |
| | Missing | 1 | 0.0% | 0 | 0.0% | 0 | 0.0% | 1 | 0.0% | 0 | 0.0% | 0 | 0.0% |
| IMD Quintiles | 1 | 72,363 | 15.4% | 28,710 | 16.7% | 284 | 21.0% | 68,334 | 14.9% | 81,965 | 18.9% | 117 | 23.0% |
| | 2 | 84,074 | 17.9% | 32,208 | 18.7% | 275 | 20.3% | 80,641 | 17.6% | 84,654 | 19.5% | 109 | 21.4% |
| | 3 | 96,109 | 20.4% | 35,481 | 20.6% | 279 | 20.6% | 93,614 | 20.5% | 85,615 | 19.7% | 102 | 20.0% |
| | 4 | 104,605 | 22.3% | 37,662 | 21.9% | 278 | 20.6% | 102,289 | 22.4% | 90,030 | 20.7% | 101 | 19.8% |
| | 5 | 111,956 | 23.8% | 37,790 | 21.9% | 232 | 15.5% | 111,706 | 24.4% | 91,371 | 21.0% | 80 | 13.6% |
| | Missing | 869 | 0.2% | 372 | 0.2% | 4 | 0.3% | 793 | 0.2% | 879 | 0.2% | 0 | 0.0% |

| | | | | | | | | | | | | | |
|---|---|---|---|---|---|---|---|---|---|---|---|---|---|
| Previously positive | No | 410,326 | 87.3% | 169,930 | 98.7% | 1,347 | 99.6% | 391,382 | 85.6% | 395,115 | 90.9% | 490 | 96.3% |
| | Yes | 59,650 | 12.7% | 2,293 | 1.3% | 5 | 0.4% | 65,995 | 14.4% | 39,399 | 9.1% | 19 | 3.7% |
| Vaccine priority groups | Healthcare worker | 18,031 | 3.8% | 4,068 | 2.4% | 4 | 0.3% | 14,693 | 3.2% | 11,743 | 2.7% | 1 | 0.2% |
| | CEV | 44,973 | 9.6% | 13,147 | 7.6% | 475 | 35.1% | 38,131 | 8.3% | 32,486 | 7.5% | 280 | 55.0% |
| | At risk** | 119,372 | 25.4% | 40,186 | 23.3% | 14 | 1.0% | 111,789 | 24.4% | 95,944 | 22.1% | 3 | 0.6% |
| | Severely immunosuppressed | 5,571 | 1.2% | 1,871 | 1.1% | 116 | 8.6% | 4,994 | 1.1% | 5,734 | 1.3% | 83 | 16.3% |

* ChAdOx1-S primary course
**At risk is only those under 65

Supplementary Table 3. Effectiveness of the ChAdOx1-S and BNT162b2 booster vaccines against hospitalisation as defined by linkage to the accident and emergency (ECDS) data following infection with Delta or Omicron variants in adults aged 65 years and older in England.

| Dose | Booster Manufacturer | Interval (days) | Controls | Cases | OR* | VE (95% CI) |
|---|---|---|---|---|---|---|
| | | | | | Delta | |
| Unvaccinated | | | 2,685 | 366 | Baseline | Baseline |
| Dose 2** | | 175+ | 21180 | 809 | 0.17 (0.15-0.2) | 82.8 (79.9 to 85.3) |
| Booster | Any*** | 0-6 | 5,502 | 96 | 0.09 (0.07-0.12) | 91.0 (88.5 to 93.0) |
| | BNT162b2 | 7+ | 64380 | 76 | 0.01 (0.01-0.02) | 98.8 (98.4 to 99.1) |
| | ChAdOx1-S | 7+ | 151 | 5 | 0.35 (0.13-0.91) | 65.3 (8.7 to 86.8) |
| | | | | | Omicron | |
| Unvaccinated | | | 1,561 | 93 | Baseline | Baseline |
| Dose 2** | | 175+ | 4329 | 84 | 0.45 (0.32-0.63) | 55.4 (37.2 to 68.4) |
| Booster | Any*** | 0-6 | 1,567 | 5 | 0.15 (0.06-0.38) | 85.2 (61.7 to 94.3) |
| | BNT162b2 | 7+ | 63803 | 320 | 0.08 (0.06-0.1) | 92.3 (89.8 to 94.2) |
| | ChAdOx1-S | 7+ | 173 | 7 | 0.34 (0.15-0.8) | 65.9 (19.9 to 85.4) |

*Odds ratio

**ChAdOx1-S primary course

***ChAdOx1-S or BNT162b2

Supplementary Table 4. Descriptive characteristics of those included in the VE analysis for protection against hospitalisation as assessed using the SUS data following infection with either Delta or Omicron variants.

| | | | | Delta | | | | | | Omicron | | | | | |
|---|---|---|---|---|---|---|---|---|---|---|---|---|---|---|---|
| | | | | Overall | | Controls | | Cases | | Overall | | Controls | | Cases | |
| | Dose | Booster manufacturer | Interval (days) | n | % | n | % | n | % | n | % | n | % | n | % |
| | | | | 23,227 | 100.0% | 17,377 | 74.8% | 5,850 | 25.2% | 11,052 | 100.0% | 9,021 | 81.6% | 2,031 | 18.4% |
| Vaccination Status | Unvaccinated | | | 3,327 | 14.3% | 1,327 | 7.6% | 2,000 | 34.2% | 1,120 | 10.1% | 517 | 5.7% | 603 | 29.7% |
| | Dose 2* | | 175+ | 10,248 | 44.1% | 7,000 | 40.3% | 3,248 | 55.52% | 2,026 | 18.3% | 1,596 | 17.7% | 430 | 21.2% |
| | Booster | Any | 0-6 | 1,366 | 5.9% | 1,123 | 6.5% | 243 | 4.15% | 388 | 3.5% | 361 | 4.0% | 27 | 1.3% |
| | Booster | BNT162b2 | 7+ | 8,239 | 35.5% | 7,884 | 45.4% | 355 | 6.07% | 7,448 | 67.4% | 6,495 | 72.0% | 953 | 46.9% |
| | Booster | ChAdOx1-S | 7+ | 47 | 0.2% | 43 | 0.2% | 4 | 0.07% | 70 | 0.6% | 52 | 0.6% | 18 | 0.9% |
| Pillar | Pillar 1 | | | 20,960 | 90.2% | 16,976 | 97.7% | 3,984 | 68.1% | 10,374 | 93.9% | 8,810 | 97.7% | 1,564 | 77.0% |
| | Pillar 2 | | | 2,267 | 9.8% | 401 | 2.3% | 1,866 | 31.9% | 678 | 6.1% | 211 | 2.3% | 467 | 23.0% |
| Gender | Female | | | 12,333 | 53.1% | 9,594 | 55.2% | 2,739 | 46.8% | 5,955 | 53.9% | 4,940 | 54.8% | 1,015 | 50.0% |
| | Male | | | 10,806 | 46.5% | 7,699 | 44.3% | 3,107 | 53.1% | 5,041 | 45.6% | 4,028 | 44.7% | 1,013 | 49.9% |
| | Missing | | | 88 | 0.4% | 84 | 0.5% | 4 | 0.1% | 56 | 0.5% | 53 | 0.6% | 3 | 0.1% |
| Age | 65-69 | | | 3,147 | 13.5% | 1,896 | 10.9% | 1,251 | 21.4% | 1,389 | 12.6% | 1,096 | 12.1% | 293 | 14.4% |
| | 70-74 | | | 4,590 | 19.8% | 3,106 | 17.9% | 1,484 | 25.4% | 2,120 | 19.2% | 1,683 | 18.7% | 437 | 21.5% |
| | 75-79 | | | 4,696 | 20.2% | 3,446 | 19.8% | 1,250 | 21.4% | 2,223 | 20.1% | 1,815 | 20.1% | 408 | 20.1% |
| | 80-84 | | | 3,514 | 15.1% | 2,691 | 15.5% | 823 | 14.1% | 1,709 | 15.5% | 1,363 | 15.1% | 346 | 17.0% |
| | 85-89 | | | 3,526 | 15.2% | 2,924 | 16.8% | 602 | 10.3% | 1,731 | 15.7% | 1,440 | 16.0% | 291 | 14.3% |
| | 90+ | | | 3,754 | 16.2% | 3,314 | 19.1% | 440 | 7.5% | 1,880 | 17.0% | 1,624 | 18.0% | 256 | 12.6% |
| Ethnicity | African | | | 127 | 0.5% | 62 | 0.4% | 65 | 1.1% | 55 | 0.5% | 23 | 0.3% | 32 | 1.6% |
| | Any other Asian background | | | 154 | 0.7% | 89 | 0.5% | 65 | 1.1% | 71 | 0.6% | 47 | 0.5% | 24 | 1.2% |
| | Any other Black background | | | 65 | 0.3% | 34 | 0.2% | 31 | 0.5% | 35 | 0.3% | 20 | 0.2% | 15 | 0.7% |
| | Any other White background | | | 1,084 | 4.7% | 746 | 4.3% | 338 | 5.8% | 490 | 4.4% | 371 | 4.1% | 119 | 5.9% |
| | Any other ethnic group | | | 199 | 0.9% | 125 | 0.7% | 74 | 1.3% | 83 | 0.8% | 58 | 0.6% | 25 | 1.2% |
| | Any other mixed background | | | 54 | 0.2% | 40 | 0.2% | 14 | 0.2% | 31 | 0.3% | 21 | 0.2% | 10 | 0.5% |

| | | | | | | | | | | | | | |
|---|---|---|---|---|---|---|---|---|---|---|---|---|---|
| | Bangladeshi or British Bangladeshi | 61 | 0.3% | 37 | 0.2% | 24 | 0.4% | 28 | 0.3% | 18 | 0.2% | 10 | 0.5% |
| | British, Mixed British | 19,165 | 82.5% | 14,768 | 85.0% | 4,397 | 75.2% | 9,137 | 82.7% | 7,647 | 84.8% | 1,490 | 73.4% |
| | Caribbean | 240 | 1.0% | 94 | 0.5% | 146 | 2.5% | 133 | 1.2% | 54 | 0.6% | 79 | 3.9% |
| | Chinese | 40 | 0.2% | 15 | 0.1% | 25 | 0.4% | 13 | 0.1% | 6 | 0.1% | 7 | 0.3% |
| | Indian or British Indian | 323 | 1.4% | 209 | 1.2% | 114 | 1.9% | 160 | 1.4% | 109 | 1.2% | 51 | 2.5% |
| | Irish | 231 | 1.0% | 185 | 1.1% | 46 | 0.8% | 123 | 1.1% | 99 | 1.1% | 24 | 1.2% |
| | Pakistani or British Pakistani | 291 | 1.3% | 169 | 1.0% | 122 | 2.1% | 148 | 1.3% | 96 | 1.1% | 52 | 2.6% |
| | White and Asian | 14 | 0.1% | 9 | 0.1% | 5 | 0.1% | 4 | 0.0% | 3 | 0.0% | 1 | 0.0% |
| | White and Black African | 10 | 0.0% | 6 | 0.0% | 4 | 0.1% | 5 | 0.0% | 5 | 0.1% | 0 | 0.0% |
| | White and Black Caribbean | 39 | 0.2% | 21 | 0.1% | 18 | 0.3% | 18 | 0.2% | 9 | 0.1% | 9 | 0.4% |
| | Missing | 1,130 | 4.9% | 768 | 4.4% | 362 | 6.2% | 518 | 4.7% | 435 | 4.8% | 83 | 4.1% |
| NHS Region | East of England | 2,549 | 11.0% | 1,997 | 11.5% | 552 | 9.4% | 1,133 | 10.3% | 972 | 10.8% | 161 | 7.9% |
| | London | 2,390 | 10.3% | 1,704 | 9.8% | 686 | 11.7% | 1,152 | 10.4% | 798 | 8.8% | 354 | 17.4% |
| | Midlands | 4,749 | 20.4% | 3,554 | 20.5% | 1,195 | 20.4% | 2,451 | 22.2% | 2,012 | 22.3% | 439 | 21.6% |
| | North East | 4,433 | 19.1% | 3,172 | 18.3% | 1,261 | 21.6% | 2,039 | 18.4% | 1,662 | 18.4% | 377 | 18.6% |
| | North West | 3,376 | 14.5% | 2,546 | 14.7% | 830 | 14.2% | 1,678 | 15.2% | 1,312 | 14.5% | 366 | 18.0% |
| | South East | 3,412 | 14.7% | 2,697 | 15.5% | 715 | 12.2% | 1,621 | 14.7% | 1,406 | 15.6% | 215 | 10.6% |
| | South West | 2,318 | 10.0% | 1,707 | 9.8% | 611 | 10.4% | 978 | 8.8% | 859 | 9.5% | 119 | 5.9% |
| IMD Quintiles | 1 | 5,443 | 23.4% | 3,913 | 22.5% | 1,530 | 26.2% | 2,727 | 24.7% | 2,095 | 23.2% | 632 | 31.1% |
| | 2 | 4,862 | 20.9% | 3,590 | 20.7% | 1,272 | 21.7% | 2,293 | 20.7% | 1,833 | 20.3% | 460 | 22.6% |
| | 3 | 4,575 | 19.7% | 3,441 | 19.8% | 1,134 | 19.4% | 2,118 | 19.2% | 1,739 | 19.3% | 379 | 18.7% |
| | 4 | 4,583 | 19.7% | 3,532 | 20.3% | 1,051 | 18.0% | 2,167 | 19.6% | 1,841 | 20.4% | 326 | 16.1% |
| | 5 | 3,718 | 16.0% | 2,873 | 16.5% | 845 | 14.4% | 1,723 | 15.6% | 1,496 | 16.6% | 227 | 11.2% |
| | Missing | 46 | 0.2% | 28 | 0.2% | 18 | 0.3% | 24 | 0.2% | 17 | 0.2% | 7 | 0.3% |
| Previously positive | No | 20,958 | 90.2% | 15,158 | 87.2% | 5,800 | 99.1% | 9,717 | 87.9% | 7,786 | 86.3% | 1,931 | 95.1% |
| | Yes | 2,269 | 9.8% | 2,219 | 12.8% | 50 | 0.9% | 1,335 | 12.1% | 1,235 | 13.7% | 100 | 4.9% |
| Vaccine priority groups | Healthcare worker | 16 | 0.1% | 12 | 0.1% | 4 | 0.1% | 6 | 0.1% | 5 | 0.1% | 1 | 0.0% |
| | CEV | 12,340 | 53.1% | 9,915 | 57.1% | 2,425 | 41.5% | 6,188 | 56.0% | 5,046 | 55.9% | 1,142 | 56.2% |

| | | | | | | | | | | | | |
|---|---|---|---|---|---|---|---|---|---|---|---|---|
| At risk* | 115 | 0.5% | 69 | 0.4% | 46 | 0.8% | 55 | 0.5% | 47 | 0.5% | 8 | 0.4% |
| Severely immunosuppressed | 1,967 | 8.5% | 1,510 | 8.7% | 457 | 7.8% | 1,022 | 9.2% | 795 | 8.8% | 227 | 11.2% |

* ChAdOx1-S primary course
**At risk is only those und

### References


1. UK Health Security Agency. COVID-19: the green book, chapter 14a. Immunisation against infectious diseases: UK Health Security Agency,; 2020.
